# Cleaner Air for Lower Cardiometabolic Risk: protocol for a double-blind, randomized, sham-controlled trial of HEPA filtration in adults with prediabetes

**DOI:** 10.64898/2026.05.29.26354420

**Authors:** Sharine Wittkopp, Parsa Asachi, Filipp Kazatsker, José O. Alemán, Terry Gordon, Robert D. Brook, Lorna E. Thorpe, Jonathan D. Newman

**Affiliations:** Leon H. Charney Division of Cardiology, NYU Grossman School of Medicine, New York, NY; Department of Medicine, NYU Grossman School of Medicine, New York, NY; Translational Research, Advent Health, Orlando FL; Department of Internal Medicine, Division of Cardiovascular Diseases, Wayne State University, Detroit, MI; Department of Population Health, NYU Grossman School of Medicine, New York, NY

## Abstract

**Introduction:** Air pollution is a leading driver of cardiovascular disease with a growing body of literature implicating this in worse glucose homeostasis. Increases in fine particulate matter air pollution (PM_2.5_) are associated with increased blood glucose and hemoglobin A1c across the glycemic spectrum from normoglycemia to prediabetes to all forms of diabetes. Despite strong evidence for positive associations of PM_2.5_ with dysglycemia, it remains unknown if reducing air pollution exposure through air filtration can effect improvements in glucose. This study aims to test the hypothesis that short-term, in-home air pollution reduction using high efficiency particulate air (HEPA) filtration will improve blood sugar in adults with prediabetes.

**Methods and analysis:** This trial is a randomized, double-blind, sham-controlled trial of the effects of lowering air pollution exposure using HEPA filtration on cardiometabolic health in adults with prediabetes living in the New York City area. Participants will be randomly assigned to use bedroom air cleaners, or sham air cleaners, while measuring PM_2.5_ continuously for 1 month. The primary outcomes will be continuous glucose monitoring metrics measured before and after HEPA air filtration. Exploratory outcomes will include insulin resistance measures, serum biomarkers and transcriptomics measured before and after HEPA intervention. We will quantify effects of HEPA filtration with models using treatment arm (true versus sham filtration) as the independent variable. Secondary analyses will model continuous measures of PM_2.5_ as the independent variable.

**Ethics and Dissemination:** This study has undergone peer review; and the work was supported by Grant 2023-0214 from the Doris Duke Foundation, who had no other role in study design or implementation. The study was registered in ClinicalTrials.gov (NCT05994937) prior to recruitment.

**Clinical Trials:** Clinical Trials NCT05994937; https://clinicaltrials.gov/study/NCT05994937

## Introduction

Despite recent improvements in air quality, more than one in five Americans are exposed to fine particulate matter air pollution (PM_2.5_, < 2.5 μm in aerodynamic diameter) concentrations exceeding US annual National Ambient Air Quality Standards [1–5]. Even exposure to PM_2.5_ below current regulatory standards (<9 μg/m^3^) has consistently been shown to promote or worsen cardiovascular risk [6–10] factors and increase mortality [11–16]. In addition, PM air pollution is implicated in the increasing prevalence of prediabetes and dysglycemia [17]. Studies suggest that increased exposure to PM_2.5_ is associated with increased blood glucose and HbA1c [18] across the glycemic spectrum, including normoglycemia, prediabetes, and all forms of diabetes [19]. *In vitro* and *in vivo* studies show additive harm of exposure to PM_2.5_ on a background of hyperglycemia [20,21], strengthening the case for a causal role for PM_2.5_ in these adverse cardiometabolic outcomes. In addition, PM_2.5_ exposure is associated with increased incidence of diabetes [22]. Results from the Global Burden of Disease study show an estimated 20% of type 2 diabetes incidence is potentially attributable to PM_2.5_ exposure [22]. Prediabetes contributes to both progression to diabetes and increased cardiovascular risk [23–27] making it an important potentially modifiable risk factor for cardiometabolic disease. Air pollution exposure, particularly to PM_2.5_ and smaller ultrafine particles, is a key environmental contributor to systemic inflammation. Exposure may therefore play an important role in the development of dysglycemia through inflammatory, as well as other mechanistic pathways, and the ensuing cardiometabolic complications [28–41]. Given that prediabetes is a common, modifiable risk factor for cardiovascular disease (CVD), understanding its drivers is key to reducing diabetes and atherosclerotic CVD and cardiometabolic risk.

Despite strong evidence that PM_2.5_ exposure is associated with impaired glucose homeostasis, it is still unclear whether lowering PM_2.5_ exposure improves glucose homeostasis metrics. Reductions in PM_2.5_ in experimental settings show beneficial effects on cardiovascular risk factors such as blood pressure and inflammatory markers associated with cardiometabolic disease [42–44]. Portable air cleaners (PACs) equipped with high-efficiency particulate air (HEPA) filters represent a practical, scalable intervention to reduce indoor PM_2.5_ exposure. HEPA filtration systems can remove ≥99% of airborne particles at 0.3 μm in diameter, substantially lowering personal exposure in indoor environments where individuals spend the majority of their time[45]. Though there are randomized-control trials investigating the effect of PACs on blood pressure [46–49], to our knowledge, there are no randomized trials that evaluate whether reducing PM_2.5_ exposure using PACs leads to improvements in glycemic outcomes, insulin resistance and other biomarkers related to cardiometabolic risk [50]. To address this gap, we propose a randomized, double-blind, sham-controlled trial to evaluate changes in glucose variability and inflammation over four weeks of PAC use. We hypothesize that four weeks of PM_2.5_ reduction using PACs, compared with sham filtration, will reduce glycemic variability as measured by continuous glucose monitoring and decrease circulating inflammatory biomarkers associated with PM_2.5_ exposure (suggesting a mechanistic linkage).

## Objectives

The primary objective of this study is to test whether reducing PM_2.5_ exposure using PACs improves glycemic control and cardiometabolic health over a four-week period in adults with prediabetes. We will evaluate the effects of home air filtration on glycemic variability, targeted and non-targeted biomarkers of cardiometabolic risk, and insulin resistance. We will test whether PAC use influences blood and urine biomarkers of air pollution exposure to understand mechanisms underlying the relationship between air pollution and glycemic control.

## Methods

### Overall study design

This “Cleaner Air for Lower Cardiometabolic Risk” study is a randomized, double-blind, sham-controlled trial, the design of which was registered prior to participant recruitment (NCT05994937). Participants are randomized in a 1:1 ratio to receive either an active PAC with true HEPA filtration or a sham PAC with the HEPA filter removed. Randomization has been computer-generated in blocks of 20 and performed by unblinded study staff not involved in outcome assessment, while all other study personnel and participants remain blinded to treatment allocation. **Figure 1** shows the schedule of enrollment, interventions and assessments for the study. The completed Standard Protocol [51] Items: Recommendations for Interventional Trials (SPIRIT) checklist is found in Supplemental information S1. A summarized study schematic is depicted in **Figure 2**.

**Figure 1.**
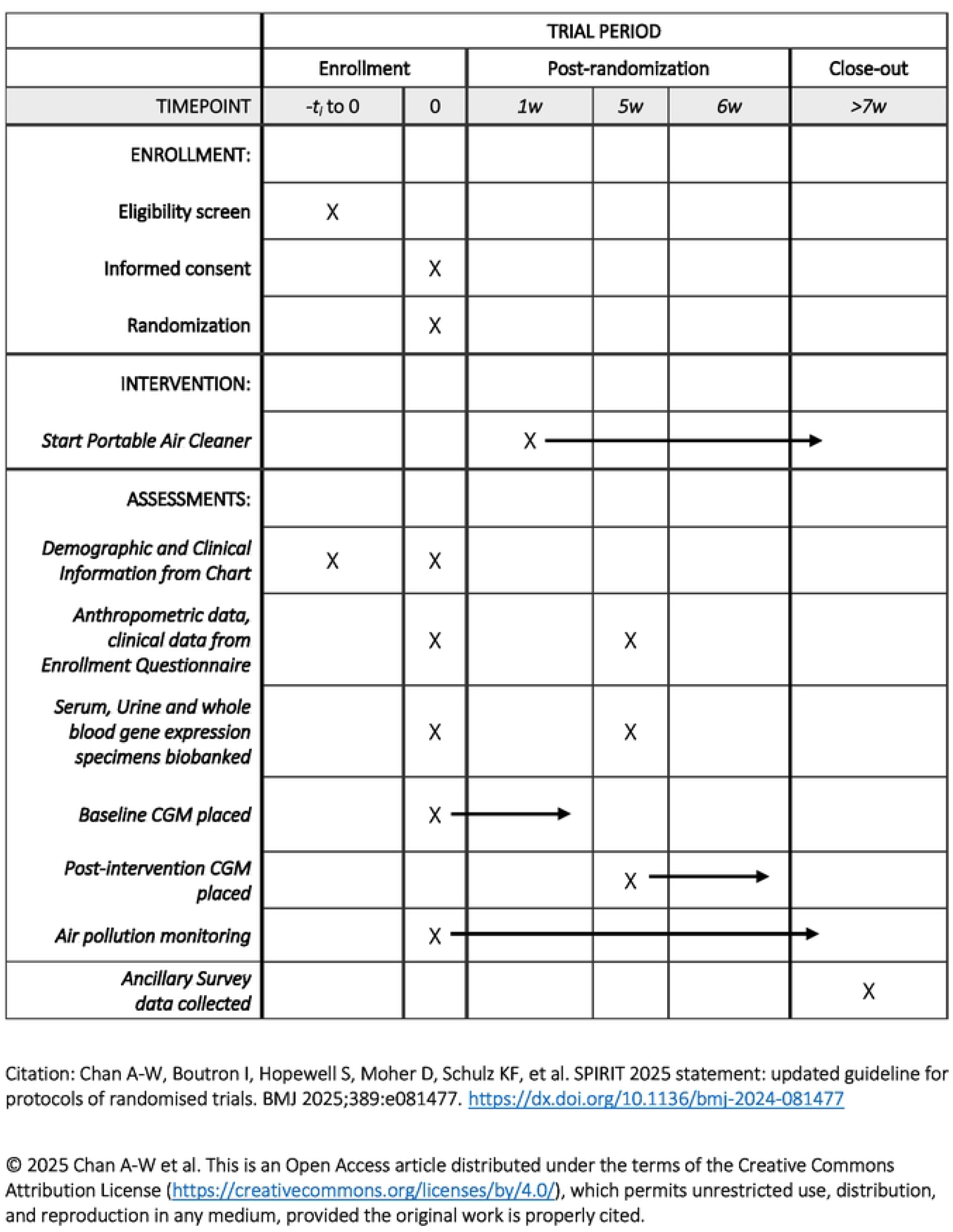
SPIRIT diagram Participant timeline: Schedule of enrollment, interventions, and assessments.

**Figure 2:**
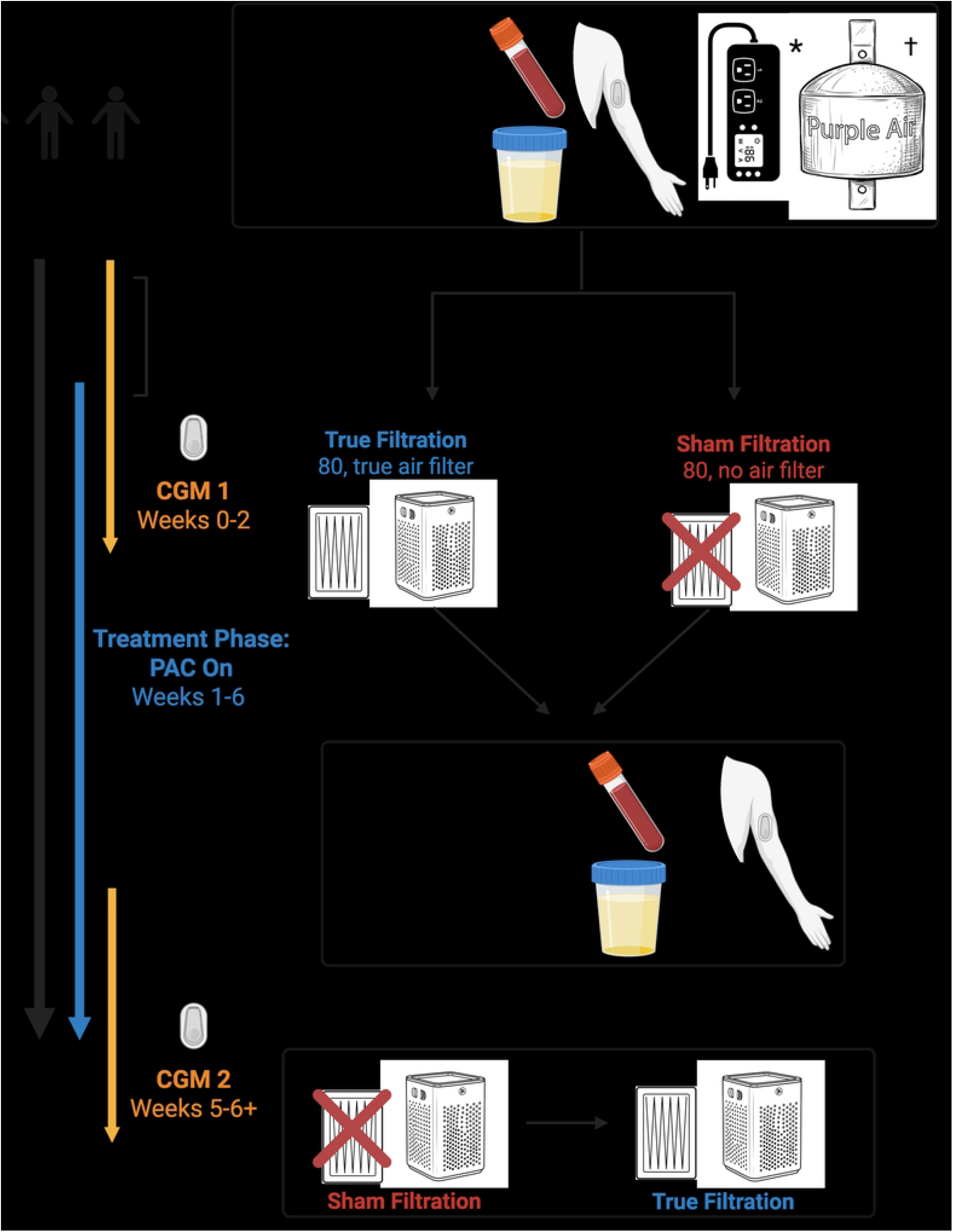
Study schematic demonstrating timing of intervention, pollution monitoring, assessments, and continuous glucose monitor (CGM) monitoring.

The primary outcome is the percent change in the continuous glucose monitoring coefficient of variation (CGM-CV) over four weeks. We will explore other dynamic glucose metrics obtained by the CGM, such as time above glucose range and mean amplitude of glycemic excursions. Additional exploratory outcomes include change in insulin resistance measured by the homeostatic model assessment of insulin resistance (HOMA-IR), change in circulating concentrations of inflammatory and cardiometabolic markers, and biomarkers of pollution exposure. Recruitment started 26 December 2023 and ended 22 April 2026; data collection is estimated to be complete by 30 July 2026 and primary results are expected 6 November 2026.

### Description of cohort and recruitment

Eligible participants include adults 18 years of age or older with a diagnosis of prediabetes (hemoglobin A1c between 5.7-6.4% within past year without a prior diagnosis of diabetes or A1c >6.4%) living in the New York City metro area. Exclusion criteria include those with first- or second-hand exposure to tobacco or other smoking or vaping in their homes and those unable or unwilling to wash-out any pre-existing, ongoing air filtration. Other specific exclusion criteria exclude those with altered blood glucose or systemic inflammation from various causes, as well as those with established atherosclerotic disease. **Table 1** lists detailed inclusion and exclusion criteria.

**Table 1.**
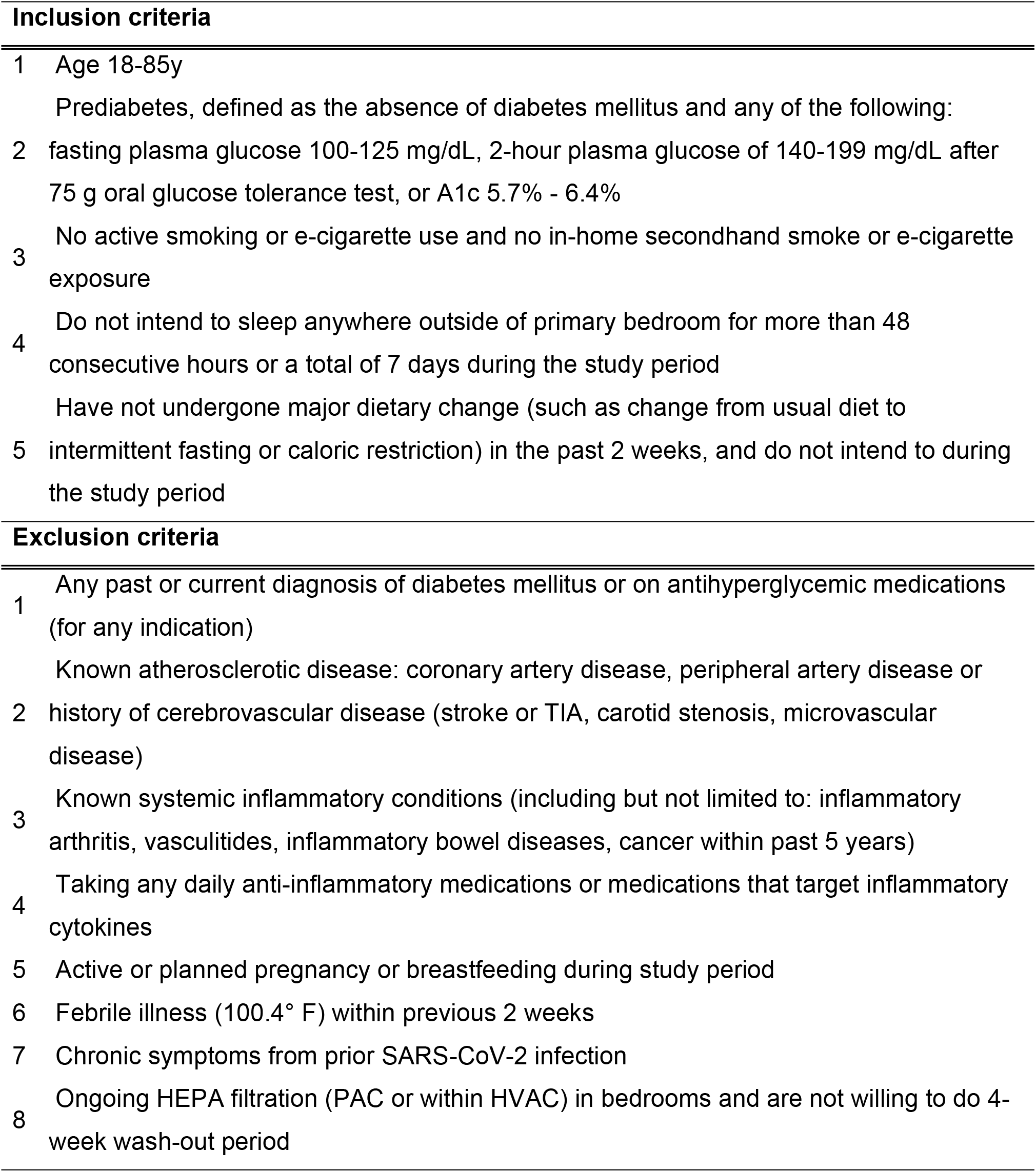
Inclusion and exclusion criteria.

Eligible participants are identified through the NYU Langone Health electronic health record using internal informatics tools. Potentially eligible participants are contacted via phone, email, or patient portal using IRB-approved scripts to confirm eligibility. Interested and eligible individuals attend research visits at NYU Langone Health Clinical and Translational Science Institute sites in Manhattan, Brooklyn, and Long Island. All enrolled participants are provided written informed consent.

### Description of intervention

The study intervention is 4 weeks of in-home PAC use. The PAC that is being used is a commercially available PAC (MedifyAir MA-25, MedifyAir, FL, USA) designed for rooms up to approximately 350 ft^2^. Although there may be substantial variability, the average primary bedroom size in the United States is approximately 250 ft^2^ and is often lower in metropolitan areas. Thus, this PAC was selected to provide optimal coverage of the air volume for nearly all participants. Because it runs continuously, it would likely still provide effective filtration for slightly larger spaces, simply taking a longer time to reach steady state [49]. For the sham arm, PACs will have HEPA filters removed and devices will be secured to prevent accidental unblinding of participants. This model of PAC has no indicator light or auto-mode which would increase the fan speed if high particulate matter is detected, thus sham devices look and sound identical to true PACs. Participants set up study equipment in their own homes using easy-to-follow, written instructions provided by study staff. Participants place the PAC in their primary sleeping quarters at least 30 cm from walls or other objects to ensure adequate airflow. Participants are instructed to operate the PAC 24 hours per day for a total of six weeks, starting one week after the initial enrollment visit (Figure 2).

### Exposure Measurement

To evaluate indoor air pollution concentration by intervention arm, we will monitor PM in participant sleeping quarters. At the initial study visit, participants receive a commercially available PM monitor placed in the same room as the PAC to record PM concentrations. The Purple Air PA-SD-II (Purple Air, Draper, UT) pollution monitor uses dual sensors to record PM concentrations on an internally housed micro-SD card every two minutes using laser-based technology. The PM_2.5_ continuous concentrations will also be evaluated as the independent variable in secondary and exploratory models. In addition to measuring PM, adherence to PAC use is quantified using electricity monitor data by comparing observed versus expected power consumption. Participants with evidence of non-use will be retained in intention-to-treat analyses.

### Outcome Measurement

At both the first and final (after week 4 of intervention) visit, trained clinical staff will place a continuous glucose monitor (CGM) (Freestyle Libre Pro, Abbott, Illinois) on the posterior upper arm using the manufacturer’s sensor applicator with standard sterile technique. Each CGM sensor stores data on interstitial glucose every 15 minutes for up to 14 days. During this time, participants are instructed not to change their diet, physical activity, or sleep habits to minimize behavior changes due to being observed. To preserve blinding, participants will not be given the CGM reader device. The CGMs are worn for two weeks each and returned by mail after completion at which time trained staff will download and store the data in a secure REDCap database. Also at each visit, we will collect venous blood and urine samples to measure serum fasting glucose and insulin, and for further biomarker evaluation. Fasting glucose and insulin will be measured by the NYU clinical laboratories according to standard protocols. Additional blood samples will be processed within two hours of collection, centrifuged, aliquoted into labeled cryovials, and stored at minus 80°C for future analysis. We will collect whole blood into PAXgene mRNA tubes (Qiagen, Hilden, Germany) and will process according to product specifications. Urine samples will be stored under identical conditions.

### Statistical Analysis

There are no data quantifying changes in continuous glucose monitoring results from air pollution reduction with PACs in any population. However, meta-analysis results showed that a 10 μg/m^3^ increase in PM_2.5_ (including daily, 3 month, and annual averages) is associated with an approximately 30 mg/dL increase in average glucose [19]. Although pooling studies of these various durations may not accurately quantify the short-term (one month) effects, we expect that reductions in PM_2.5_ will result in comparable decreases in CGM-CV. Because the intervention primarily reduces exposure during sleeping hours and the study population has baseline elevated fasting glucose, a conservative 10% effect size is estimated. Using a multiple regression model incorporating age, body mass index, gender, and treatment assignment, a sample size of 114 provides greater than 80% power to reject the null hypothesis at a significance level of 0.05. The enrollment goal of 160 participants allows for approximately 25% attrition per arm while maintaining adequate power.

Descriptive statistics will be reported as means and standard deviations or medians and interquartile ranges for continuous variables, and as counts and proportions for categorical variables. Statistical comparisons between the PAC and sham groups will be performed using a two-group two-sided t-test or nonparametric Wilcoxon test for continuous variables and Chi-square test for categorical variables. Distributions of CGM-CV will be examined, and non-normally distributed dependent variables (outcomes) will be log-transformed, as needed.

The primary outcome will be analyzed using two complementary approaches. First, we will use a linear model assessing the change in CGM-CV from pre-to post-intervention as an effect of study arm (true versus sham), for participants with complete paired data. Second, we will fit a linear mixed-effects model using all available CGM-CV measurements, with time, study arm, and their interaction as fixed effects. This approach allows inclusion of participants with incomplete data and accounts for within-subject correlation over time. This process will be repeated for any exploratory analyses including Time Above Range (TAR, 140-180 mg/dL) and overnight CV. These models use subject-specific random intercepts to account for heterogeneity between subjects and will account for within subject correlation between biomarker observations. The proposed model is:

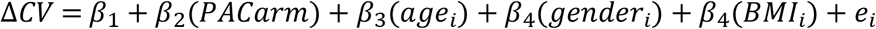

We will also utilize similar models incorporating PM_2.5_ as the independent variable in place of PAC arm. This will be assessed as the difference between pre-intervention baseline PM_2.5_ concentration and the time-weighted average concentration of PM_2.5_ from the 14-day intervention period. For our exploratory aims, we will evaluate the associations of PM_2.5_ concentration changes with change in HOMA-IR as the dependent variable. We will also explore the cardiometabolic biomarker panel results using mixed-effects models for the remaining biomarkers.

### Data management

All quantitative data will be stored in REDCap within encrypted, access-controlled databases. Biospecimens will be labeled with unique identifiers and stored at ™80°C in secure laboratory facilities. The final dataset will be retained by the investigative team and de-identified for analysis.

### Risk of Participation

This is a minimal-risk study. PACs and pollution monitors are commercially available devices with no known significant risks. Adverse events related to continuous glucose monitoring or biospecimen collection will be monitored at participant visits. There are no predefined stopping rules for either efficacy or futility.

## Conclusion

This protocol describes a double-blind, randomized, sham-controlled, pragmatic trial evaluating the cardiometabolic effects of reducing personal air pollution exposure in adults with prediabetes. The findings will address a critical gap in the field by testing whether personal reductions in PM_2.5_ exposure using portable air cleaners lead to meaningful improvements in dysglycemia and reductions in biomarkers widely associated with cardiometabolic disease in a population at elevated risk.

Epidemiological and *in vitro* studies have established associations between increased particulate matter exposure and an increased risk of diabetes. However, a causal relationship has not yet been definitively established. No randomized controlled studies thus far have directly investigated whether reducing personal exposure to PM_2.5_ alters glycemic parameters. By experimentally reducing PM_2.5_ exposure using portable air cleaners, we aim to strengthen causal inference by demonstrating biological plausibility and a dose–response relationship between personal PM_2.5_ reduction and metabolic improvement. This study represents a necessary step translating robust epidemiological associations into interventional human evidence. The results of this study seek to position PM_2.5_ exposure not only as a societal concern, but also as a modifiable, patient-level risk factor that health care providers can monitor and address. Moreover, these findings may lay the groundwork for considering PAC use as a potential component of diabetes and cardiovascular disease prevention initiatives.

A key and novel feature of this study is the use of continuous glucose monitoring to assess glycemic variability, which captures dynamic fluctuations in glucose that are not detectable using conventional measures such as fasting glucose or HbA1c and may be more sensitive to short-term environmental exposures. In addition, granular glucose measurements over the course of the study will help elucidate temporal relationships between exposure reduction and glycemic effects, enhancing mechanistic insight. The measurement of cardiometabolic biomarkers before and after PM_2.5_ reduction may also delineate which biomarkers, and thus which biological pathways, are most responsive to air pollution reduction, providing crucial effect size estimates to inform the design of larger trials. Additionally, examining changes in systemic inflammatory markers, endothelial dysfunction markers, lipid fractions, and measures of insulin resistance may help clarify the intermediate pathways through which PM_2.5_ exposure influences cardiometabolic risk.

In a broader context, demonstrating clinically meaningful reductions in cardiometabolic risk through personal air pollution reduction may have important implications for health policy and air quality regulation, particularly as millions of individuals remain exposed to unsafe levels of particulate matter.

## Data Availability

The de-identified participant data from the final research dataset used in the published manuscript will be shared upon reasonable request beginning 9 months and ending 36 months following article publication or as required by a condition of awards and agreements supporting the research provided the investigator who proposes to use the data executes a data use agreement with NYU Langone Health. Requests may be directed to. The protocol and statistical analysis plan will be made available on Clinicaltrials.gov only as required by federal regulation or as a condition of awards and agreements supporting the research

## Acknowledgements

This study was supported by Doris Duke Charitable Foundation… (They have some specific wording, I will look up and add)

